# New Admissions and Asymptomatic TB Cases fuel TB Epidemic in Prisons, a Cross Sectional Survey in Tanzania

**DOI:** 10.1101/2023.08.23.23294476

**Authors:** Chacha David Mangu, Petra Clowes, Jan van den Hombergh, Clement Mwakabenga, Simeon Mwanyonga, Jane Ambindwile, Faith Kayombo, Monica Minja, Samuel Kalluvya, Lisa Gerwing-Adima, Christa Kasang, Andreas Mueller, Edward Chilolo, Juma Angolwisye, Dickson Nsajigwa, Adili Kachima, Deus Kamala, Beatrice Mutayobya, Nyanda Elias Ntinginya, Michael Hoelscher, Elmar Saathoff, Andrea Rachow

**Affiliations:** National Institute for Medical Research, Mbeya Medical Research Center, Tanzania; PharmAccess International, Tanzania; Bugando Medical Centre, Mwanza, Tanzania; Medical Mission Institute, Wuerzburg, Germany; Ministry of Home Affairs; Prison Authority; National TB and Leprosy Program, Tanzania; Division of Infectious Diseases and Tropical Medicine, University Hospital, LMU Munich, Germany; German Center for Infection Research (DZIF), Partner Site Munich, Germany

## Abstract

**Background:** There is an increased risk for tuberculosis (TB) infection and disease progression in prison settings. TB prevalence in prisons of high and middle/low income countries have been measured to be between 5 and 50 -times higher than in the general population.

**Methods:** We performed TB active case finding in five central prisons, Keko, Segerea, Ukonga, Butimba and Ruanda prison in Tanzania, using the Xpert MTB/RIF^®^ assay on early morning or spot sputum sample from inmates and new entries between April 2014 and July 2015.

**Results:** Out of 13,868 prisoners tested, 13,763 had valid results. TB prevalence was 1.55% (214/13,763); new admissions contributed to the majority (61.68%) of TB cases, but prevalence was higher among inmates (1.75%) compared to new admissions (1.45%). Ukonga, an urban prison which incarcerates long-term convicted inmates had the highest prevalence of 4.02%. Male gender (OR=2.51, p<0.001), repeated incarcerations (OR=2.85, p<0.001), history of TB treatment (OR=1.78, p =0.002), TB symptoms (OR=2.78, p=0.006) and HIV infection (OR=2.86, p=0.002) were associated with positive TB results.

**Conclusion:** New admissions could be the driving force of the TB epidemic in the penitentiary system. However, prison environments remain a major risk factor for developing active TB disease.

## INTRODUCTION

Despite the efforts to reduce the global burden of TB, it has been one of the infectious diseases causing substantial morbidity and mortality especially in Asia and Sub-Saharan Africa^1, 2^. It is estimated that 10.6 million people developed TB in 2021, 25% of which occurred in Africa and caused 1.6 million deaths^3^. TB remains one of the major causes of death among people living with HIV (PLHIV) causing 187,000 deaths in 2021 ^3^. TB cases are still under-reported; approximately 6.4 million cases (only 60.4% of global estimated cases) were diagnosed and notified in 2021^3^. The majority of missed TB cases most likely occur among people who lack free or direct access to health care services such as prisoners and people incarcerated. Worldwide, it is estimated that more than 9 million people are incarcerated at any point in time^2^, and therefore faced with a high risk environment for new TB infection and fastening the progression of latent to active TB which, among others, is promoted by overcrowding, poor ventilation and poor diet ^4, 5^. Limited access to health care services results in late case detection and delayed treatment initiation with potential poor adherence sustaining the ongoing TB transmission in prisons ^4, 5^.

Despite its highly endemic nature, prevalence of TB in prisons is largely unknown in many countries. Systematic reviews showed incidences of 237.6 per 100,000 and 1,942.8 per 100,000 person-years in high and middle/low-income countries, respectively^6, 7^. Some studies have shown TB rates among inmates as high as 5 to 50 times greater than those of the general population across developed and developing countries, respectively^8^. Active mass screening studies in Malawi, Ivory Coast, Zambia, Botswana, Cameroon and Ethiopia, found TB prevalence among prisoners of about 7 to more than 10 times higher than in the general population^9-14^. In Tanzania, a study at Bugando Hospital, Mwanza, showed a proportion of 41% of smear-positive TB cases among prisoners with presumptive TB referred for diagnosis^15^.

Despite the high TB prevalence and complexity that exists in the control of tuberculosis in prisons, TB detection in most prisons still rely on symptomatic screening^16^, however with the availability of approved molecular diagnostic tests, such as GeneXpert, which are more sensitive, easy to perform and rapid, the systematic TB screening in a large number of persons within a short time has been made possible^19^. The aim of this study was to implement a screening strategy to actively detect TB in five central prisons in Tanzania mainland using GeneXpert MTB/RIF and accurately measure the prevalence, and understand the related risk factors of TB in prisons.

## METHODS

### Prisons and study groups

The project was implemented in five central prisons in Tanzania; Keko, Segerea and Ukonga Prisons within the highly populated business capital of the Indian Ocean coast region Dar es Salaam; Butimba Prison in Northwest region, Mwanza; and Ruanda Prison in the Southern Highland region, Mbeya. All prisons are located in urban settings but admit prisoners from both urban and rural communities. According to the Tanzanian prison authority, during the study period, these prisons housed an average of about 8,000 inmates in total and have a turnover of more than 15,000 new admissions and discharges per year. TB screening was done among those who newly entered the prison, were transferred in from another prison and inmates present in the prisons at the beginning of the study.

### Study procedures

The project was implemented between April 2014 and June 2015. A screening questionnaire administered by trained prison health staffs. was used to collect demographic information, date of entry into prison, history of previous imprisonment, number of inmates sharing a cell, presence of cell mate(s) treated for TB, TB symptoms, self-reported past history of TB treatment, self-reported HIV status and use of anti-retroviral treatment (ART). One spontaneously expectorate sputum sample was collected from all study participants by a trained prison health facility staff, a nurse or laboratory technician. TB diagnosis was done using the GeneXpert MTB/RIF^®^ assay (Xpert; Cepheid, Sunnyvale, California, US), which was stationed at each prison health facility and operated by trained laboratory technicians who were also prison staff. In case of a failed test (“error”, “invalid” or “no result”), Xpert testing was repeated on the same sample or a newly collected sample if the sample was either inadequate, discarded or of poor quality. For those consented for HIV testing, HIV diagnosis was done according to the national guidelines using SD Bioline HIV-1/2 3.0 (Standard Diagnostics, Inc. Yongin, South Korea), and confirmed by Alere Determine HIV-1/2 (Alere Inc. Massachusetts, USA) and Uni-Gold HIV-1/2 (Trinity Biotech, Co. Wicklow, Ireland).

### Ethical consideration

The study obtained ethical clearance from the National Health Research and Ethics Committee of the National Institute for Medical Research, Tanzania. It was also approved by the Prison Headquarters as well as the National TB and Leprosy Program in Tanzania.

Screening was voluntary; prisoners consented to participate and thumb printed the informed consent form before study procedures were done. HIV diagnosis was done through provider initiated testing and counseling (PITC) by a nurse counselor. Prisoners who refused to test for HIV were not excluded from TB screening. Those diagnosed to have TB or HIV were linked to care at the respective prison health facilities. Only prisoners 18 years and above were included in the study.

### Statistical analysis

The main endpoint of this study was bacteriologically confirmed TB diagnosed by Xpert. Univariate analysis was performed to describe the study population of screened prisoners and the prevalence of TB in different strata of the independent variables.

A conceptual frame work (Fig. 1) was established to guide multivariable logistic regression analysis in three levels. Variables with a p-value of <0.1 were retained and used to adjust for variables in the subsequent level and retained in the last model; and variables in the last model with the p-value of <0.05 were considered significant risk factors for TB in prisons. Variables considered distal to the outcome and those proximal to the outcome were in the first and last levels respectively. Number of cell mates was forced into the final model due to its importance in assessing overcrowding within prison cells. Robust standard errors were used in logistic regression to account for correlation within prisons. The Wald test was used to assess significance of the contribution of each variable to the model. Data analysis was performed using Stata/SE version 14 (Stata Corporation, College Station, Texas, US).

**Figure 1.**
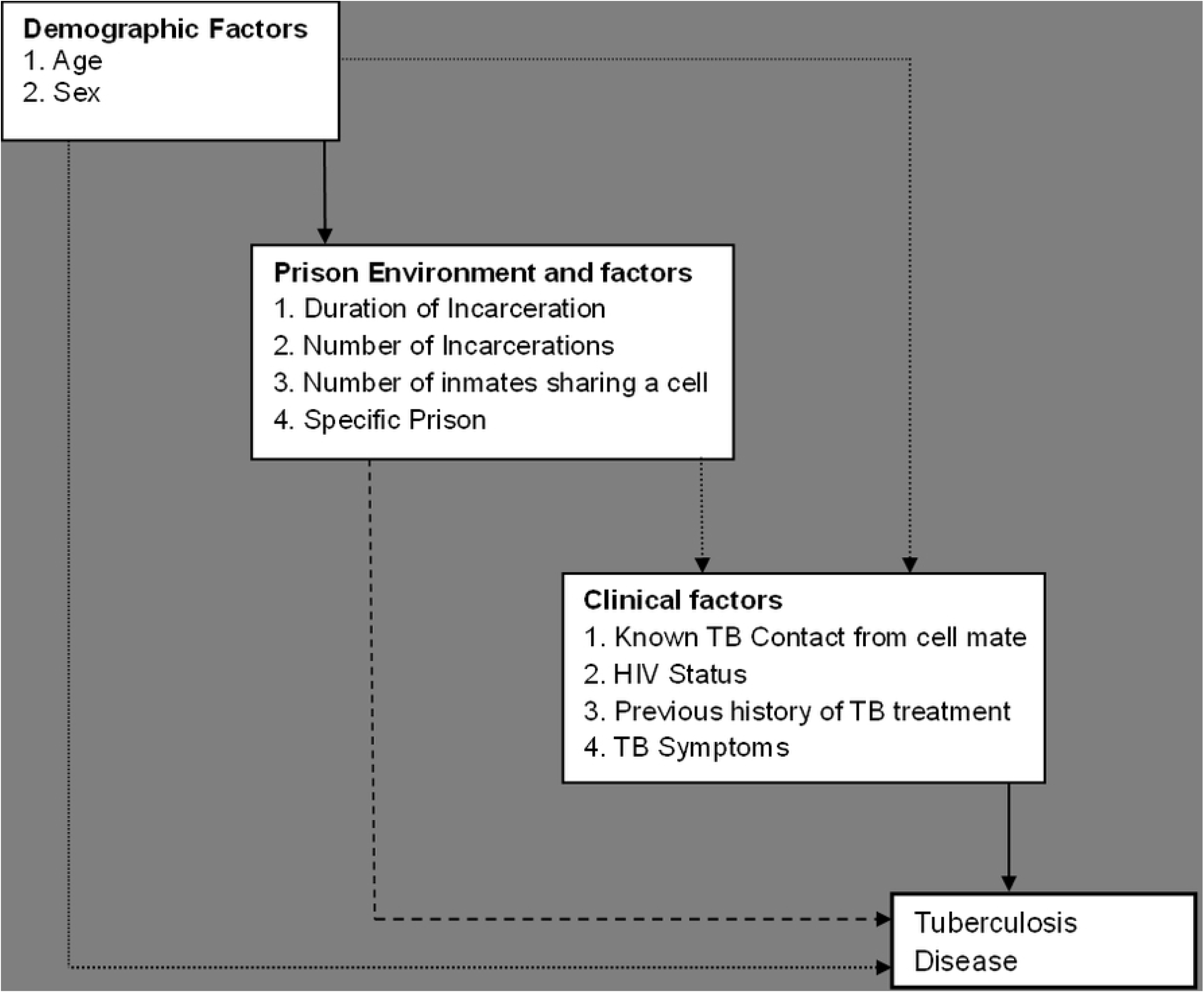
Hypothesized conceptual framework to guide model building for multivariate analysis.

## RESULTS

A total of 16,132 prisoners from 5 central prisons agreed to participate in the study and completed the questionnaire. Of these, 13,868 prisoners were screened using Xpert, of which 105 had either an invalid or erroneous final result. Thus, the final analysis included 13,763 prisoners with valid Xpert result.

### Baseline characteristics

Of the 13,763 prisoners, majority (96.3%, n=13,248) were male. The median age of screened prisoners was 30 years (IQR 24–36 years) while 64.7% were younger than 35 years. A total of 9,077 (66.0%) prisoners screened were new admissions and 12,035 (87.4%) were in prison for the first time. The median duration of incarceration was 1.5 months at the time point of study participation and sharing a cell with more than 10 inmates was common (93.8%, n=12,911). More than half (51.6%, n=7,106) had unknown HIV status, and 6.1% (n=833) were HIV positive (Table 1).

**TABLE 1.** Characteristics of the study population and TB cases.

| Characteristics | Categories | Total<br>(col%)<br>[N=13,763] | Prisoners with<br>tuberculosis<br>(col%)<br>[N=214] | Prevalence of<br>Tuberculosis<br>in % | P-value* |
| --- | --- | --- | --- | --- | --- |
| Sex | Female | 515 ( 3.7) | 4 (1.9) | 0.78 | 0.150 |
|  | Male | 13,248 (96.3) | 210 (98.1) | 1.59 |  |
| Age in years | <25 | 3,731 (27.1) | 35 (16.4) | 0.94 | 0.004 |
|  | 25-34 | 5,177 (37.6) | 87 (40.7) | 1.68 |  |
|  | 35-44 | 3,030 (22.0) | 58 (27.1) | 1.91 |  |
|  | >45 | 1,825 (13.3) | 34 (15.9) | 1.86 |  |
| Prisoner Status | New Admission | 9,077 (66.0) | 132 (61.7) | 1.45 | 0.184 |
|  | Inmate | 4,686 (34.0) | 82 (38.3) | 1.75 |  |
| Duration of<br>Incarceration | <6 months | 8,643 (62.8) | 114 (53.3) | 1.32 | 0.023 |
|  | 6-24 months | 1,703 (12.4) | 37 (17.3) | 2.17 |  |
|  | 24-60 months | 1,246 (9.1) | 21 (9.8) | 1.69 |  |
|  | >60 months | 2,171 (15.8) | 42 (19.6) | 1.93 |  |
| Number of<br>Incarcerations | 1 | 12,035 (87.4) | 163 (76.2) | 1.35 | <0.001 |
|  | 2-3 | 1,573 (11.4) | 41 (19.2) | 2.61 |  |
|  | ≥4 | 155 (1.1) | 10 (4.7) | 6.45 |  |
| Number of Cell Mates | <5 | 47 (0.3) | 2 (0.9) | 4.26 | 0.195 |
|  | 6-10 | 805 (5.9) | 9 (4.2) | 1.12 |  |
|  | >10 | 2,911 (93.8) | 203 (94.9) | 1.57 |  |
| Known TB contact<br>from Cell Mate | No | 13,713 (99.6) | 212 (99.1) | 1.55 | 0.162 |
|  | Yes | 50 (0.4) | 2 (0.9) | 4.00 |  |
| Previously treated for<br>TB | Yes | 666 (4.8) | 33 (15.4) | 4.95 | <0.001 |
|  | No | 13,097 (95.2) | 181 (84.6) | 1.38 |  |
| Reported one or more<br>TB symptoms | Yes | 2,261 (16.4) | 92 (43.0) | 4.07 | <0.001 |
|  | No | 11,502 (83.6) | 122 (57.0) | 1.06 |  |
| HIV Status | Negative | 5,824 (42.3) | 85 (39.7) | 1.46 | <0.001 |
|  | Positive | 833 (6.1) | 40 (18.7) | 4.80 |  |
|  | Not Known | 7,106 (51.6) | 89 (41.6) | 1.25 |  |
| Prison ID | Ruanda | 4520 (32.8) | 42 (19.6) | 0.98 | <0.001 |
|  | Butimba | 4328 (31.5) | 48 (22.4) | 1.12 |  |
|  | Ukonga | 828 (6.0) | 32 (15.0) | 4.02 |  |
|  | Keko | 1921 (14.0) | 26 (12.2) | 1.37 |  |
|  | Segerea | 2166 (15.7) | 66 (30.8) | 0.03 |  |

\*chi squared test p-value for difference in TB-prevalence between strata

### Risk factors associated with TB diagnosis

In multivariable analysis, males had more than two times the odds for TB infection than females (Odds Ratio [OR] = 2.51, 95% confidence interval [CI] = 1.54 to 4.08, p-value [p] = <0.001). Age did not appear to be an important independent risk factor for active TB. The OR significantly increased with the frequency of incarcerations to nearly three times higher odd for active TB in prisoners who have been in prison four or more times (OR=2.85 CI=1.78 to 4.55, p<0.001). Previous history of TB treatment was associated with current active TB disease (OR=1.78, CI=1.24 to 2.55, p=0.002). Participants with TB symptoms were three times more likely to test positive for TB than those without symptoms (OR=2.78, CI=1.35 to 5.74, p<0.006) and HIV positive prisoners also had significantly higher odds of TB than HIV-negative prisoners (OR=2.86, CI=1.49 to 5.52, p=0.002). Being incarcerated in prisons located in the business capital, Ukonga prison (which admits convicted offenders) or Segerea prison (which admits new remandees) was associated with a two times higher OR for having TB (OR= 2.15, CI 1.63 to 2.83, p<0.001 and OR=2.10, CI 1.75 to 2.52, p<0.001 respectively). Sharing a cell with more than 10 people was not associated with TB infection (Table 2).

**TABLE 2.**
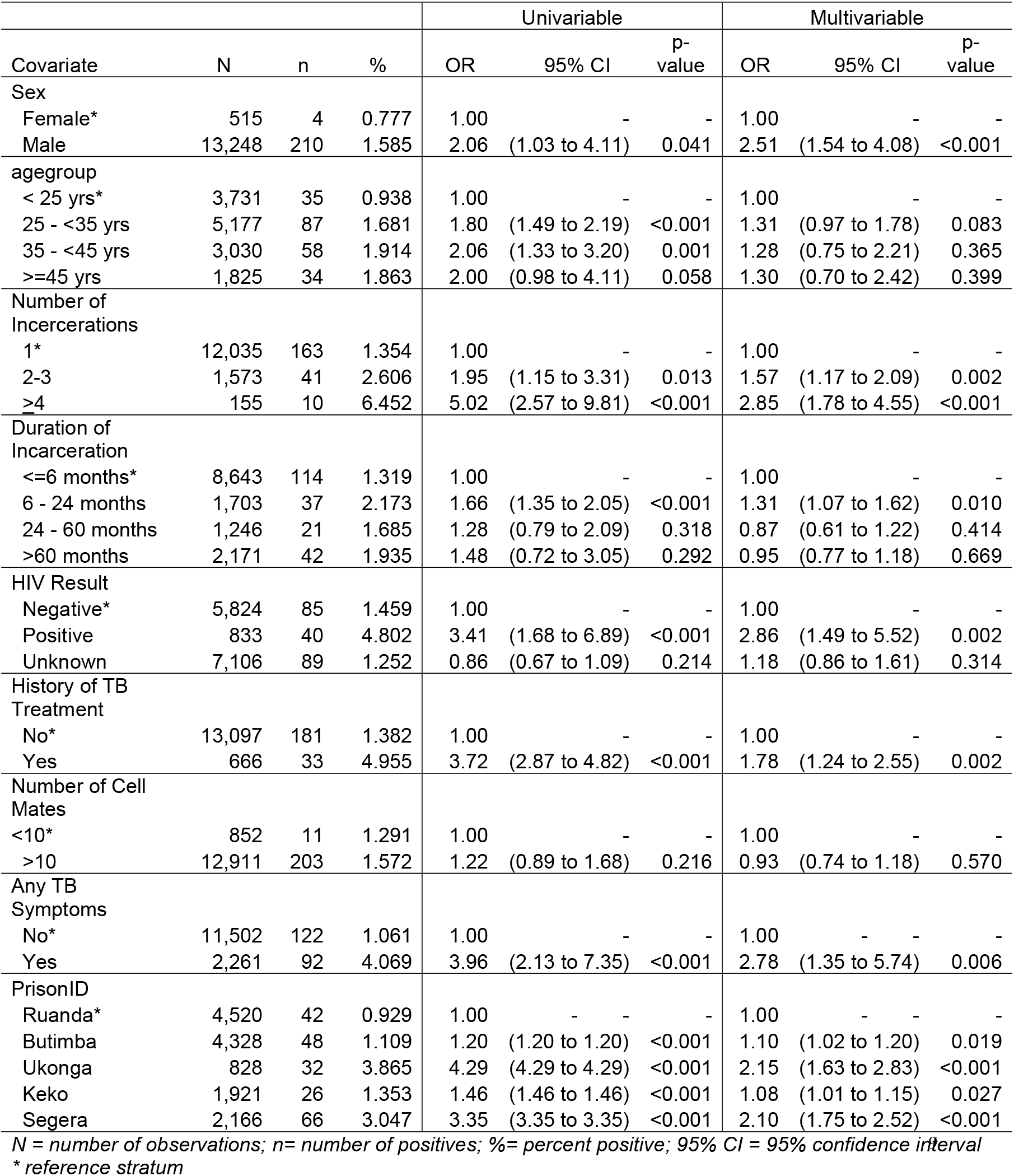
Association of various factors with TB disease (N = 13,763; n= 214 (1.555%): Uni- and multi-variable logistic regression results; using robust variance estimates adjusted for clustering by PrisonID.

## DISCUSSION

Our findings show a high TB burden in Tanzania prisons. The prevalence of bacteriologically confirmed TB of 1.55% (1,550 cases/100,000 population) is more than 5 times higher than the prevalence in the general population of 295 per 100,000^18^.

Our study contributes important findings over the containment of asymptomatic TB cases in prison settings. Sustained and prolonged exposure to TB bacilli could be maintained in prison walls by asymptomatic cases and can potentially propagate TB transmission among inmates if conventional symptomatic TB screening is the only screening method relied on since studies have shown that asymptomatic TB cases have the ability to infect others^16,19-20^. For this reason robust and effective approaches using effective diagnostic tools for TB screening in prisoners are required. Nonetheless, TB symptoms remain a major predictor of TB disease as shown by a TB high prevalence in prisoners with symptoms compared to those without (4.07% vs 1.06% respectively). Therefore an effective screening intervention is the one that apply mixed methods that detect both symptomatic and asymptomatic TB.

Despite high TB prevalence among inmates, the fact that 61% of detected TB cases in this study were among new admissions suggests that new admissions bring TB into the prison facilities and therefore contribute significantly to the TB epidemic in prisons. These findings also observed in other studies^21, 22^ in countries with high TB burden suggests two possible explanations; firstly, many of those who commit offenses and end up in a penal institution come from a background of high risk for TB infection and therefore bring with them an increased risk for TB infection into prison and actively contribute to the high prevalence of TB in prisons^5^. And secondly, it is highly likely that specific conditions in prisons lead to a higher risk of TB infection and accelerate latent TB infections to progress to active disease within a short duration of time. Many TB control strategies in prisons focus on already incarcerated inmates while little or no attention is given to new admissions. Findings from our study suggest that equal weight has to be given to both groups of prisoners. Constant screening at prison-entry aims at detecting untreated admissions and thereby reduce introduction of new TB cases into prison while mass and contact screening should aim at detecting the circulating TB within the prison walls^23^.

Our findings do not suggest overcrowding to be a risk factor for TB disease in prison; however, this might be due to the possibility that the disaggregation by number of inmates per cell during data collection was not specific enough and hence it probably obscures the association. Similarly, having a known TB contact within the cells was not associated with TB infection in our study, however, this finding could be due to the fact that more than 50% of the identified TB cases were new entries who didn’t have prolonged contact with other inmates during the time of screening. Yet, screening of individuals who have contact with index cases should still continue as recommended because it is effective in increasing case detection^23, 24, 25^. HIV remains an important single predictor of TB infection in prison settings as it is well known to fuel the TB epidemic^26^. Notably, effective control of TB infection also requires appropriate interventions against HIV infection.

## CONCLUSION AND RECOMMENDATIONS

Our study showed a high TB burden in Tanzanian prisons. The majority of positive TB tests was among symptomatic prisoners, new entries and inmates who stayed only for a short time in prison. Other TB risk factors were not prison-associated. Prisoners remain a high risk population with great mobility due to perpetuating referrals between prisons and interactions with the outside communities; our results suggest that TB screening measures within prisons need to be sensitive, robust and with a fast enough turn-over time to combat TB in prisoners and consequently interrupt TB transmission not only in prisons but also in the surrounding communities.

TB screening on admission or exit for both remandees and inmates using WHO recommended PCR based rapid diagnostics such as GeneXpert was not a routine practice in Tanzania prisons at the time of this study. However, because of the findings of this study, active TB screening on all prison entry points have been implemented in Tanzania ever since. Future research is needed to assess the impact of the implementation of WHO approved rapid TB diagnostic tools in prison settings.

## Data Availability

The data can be made available only with permission form the Tanzania Ministry of Home Affairs and Prison Authority and other local regulatory bodies due to the sensitivity of the prison data.

## Acknowledgements

We acknowledge the effort from all collaborating institutions implementing the study, NIMR-Mbeya Centre, Tanzania (project lead); PharmAccess International, Tanzania; Medical Mission Institute of Wuerzburg, Germany; Bugando Referral Hospital, Tanzania; Division of Infectious Diseases and Tropical Medicine, University of Munich (LMU) Germany (grant holder). We thank the Tanzania Prison Authority Headquarter, and the National TB and Leprosy Program for facilitating study activities. To all the prison health staffs for your dedication and prisoners for your participation, we thank you.

## Authors Contribution

CDM, PC, MH, AR designed the study, applied for grant, supervised study implementation, and reviewed the manuscript.

CDM, ES performed data analysis.

JH, CM, JA, FK, MM, SK, LG, CK, AM, DK, & NE reviewed the study design, coordinated and supervised study implementation, and reviewed the manuscript;

CDM wrote the manuscript

